# Effects of Glutamine Supplementation Combined with Exercise on Isokinetic Performance and Biochemical Parameters After Anterior Cruciate Ligament (ACL) Reconstruction: A Randomized Controlled Trial

**DOI:** 10.64898/2026.01.13.26343358

**Authors:** Hernández-Valencia Sandra Elvia, Moreno-Altamirano Laura, León-Ballesteros Saul, Salas-Romero Rebeca, Pegueros-Pérez Andrea, Carrillo-Medina Santiago Alejandro, Mendoza-Gutierrez Jonatan, Jorgan Jared Pérez Varela, Ernesto Roldan-Valadez

**Author notes:** Correspondign autor: Hernández-Valencia Sandra Elvia, B.Sc, M.Sc, PhD.

## Abstract

**Objective:** To evaluate the effects of glutamine supplementation combined with an exercise program on isokinetic performance, biochemical parameters, and muscle asymmetry indices in individuals undergoing rehabilitation following ACL reconstruction.

**Design:** A 6-week randomized controlled trial was conducted, assigning participants to either a glutamine supplementation group (ExGln) or a placebo group (ExPla), both undergoing a standardized exercise program.

**Setting:** The study was performed in a sports medicine facility with access to isokinetic testing and biochemical analysis.

**Participants:** A total of 30 participants who had undergone ACL reconstruction surgery were included, with 15 in each group.

**Methods:** Peak torque and muscle asymmetry of knee extensors and flexors were assessed via isokinetic testing. Biochemical analyses (glucose, lipid profile, liver enzymes) were conducted pre– and post-intervention. Wilcoxon and Mann–Whitney U tests were used for intra– and intergroup comparisons, respectively.

**Results:** The ExGln group showed significant improvements in peak torque for the involved limb extensors (p=0.001) and flexors (p=0.001), as well as reductions in extensor and flexor asymmetry indices (p=0.001 and p=0.018, respectively). Additionally, significant reductions were observed in C-LDL, C-HDL, and triglycerides levels in the ExGln group (p=0.006, p=0.003, and p=0.003, respectively). The intergroup analysis indicated significant differences in alkaline phosphatase levels (p=0.031). No adverse effects were reported.

**Conclusion:** Glutamine supplementation combined with a structured exercise program significantly enhanced isokinetic performance and improved biochemical parameters, suggesting a beneficial role in postoperative rehabilitation following ACL reconstruction. Further studies with larger sample sizes are recommended to validate these findings.

## Introduction

The anterior cruciate ligament (ACL) plays a critical role for knee stability and kinematics. ACL injuries, partial or complete, are common orthopedic conditions, with 100,000 to 200,000 cases annually in the United States and increasing worldwide (1, 2). These injuries may cause functional impairments, pain, and long-term complications including meniscal damage, cartilage degeneration, and osteoarthritis, developing in up to 80% of untreated cases (3, 4). Untreated ACL injuries and even surgical interventions are associated with a high risk of osteoarthritis (OR= 6.81, 95% CI 5.7-8.13, p=<0.0001 for untreated; OR= 7.7, 95% CI 6.05-9.79, p= 0.0001 for surgical treatment) (5, 6).

Despite advancements in surgical techniques, anterior cruciate ligament reconstruction (ACLR) often results in persistent deficits in quadriceps strength, affecting more than 60% of patients for up to two years postoperatively (7, 8). Quadriceps strength is essential for knee stability, functional recovery, and return to activity. Weakness has been linked to altered gait patterns, decreased physical performance, and increase risk of ACL reinjuries or falls (9, 10). Consequently, rehabilitation programs emphasizing strength training are considered the cornerstone of post-ACLR recovery. These programs aim to restore muscle mass and functional symmetry between the operated and contralateral limb; however, achieving optimal outcomes remains challenging and often prolonged (11, 12).

Nutritional supplementation, particularly with amino acids such as glutamine (Gln), may serve as a useful adjunct to standard rehabilitation protocols. Glutamine, the most abundant non-essential amino acid in the body, comprises over half of the free amino acid pool in skeletal muscle and contributes to protein synthesis and muscle recovery (13, 14). Glutamine also supports immune function and antioxidant defense, both critical during catabolic stress after ACL injury. Supplementation may promote protein synthesis, limit muscle loss and aid recovery, particularly in catabolic conditions (15–19). However, evidence remains inconsistent and inconclusive, specially regarding its efficacy on quadriceps strength and symmetry following ACLR (20–24).

This study evaluates the efficacy of oral glutamine supplementation combined with a standardized strength exercise program in adult males undergoing ACLR. The findings aim to inform evidence-based recommendations to optimize post-ACLR rehabilitation, particularly in resource-limited settings such as Mexico, where comprehensive rehabilitation tools may not be widely available (25). The ultimate goal is to enhance clinical decision-making and improve outcomes for patients with ACL injuries.

## Methods

This was a randomized, double-blind, controlled trial. Participants were randomized into two parallel groups with 1:1 ratio. The trial complied with Consolidated Standards of Reporting Trials guidelines (26). ***ClinicalTrials.gov*** (NCT03517254).

### Participants

Male aged 20-50 years, 10-15 weeks post ACLR, with moderate quadriceps strength deficits (R H/Q: 70-74.9%) on isokinetic testing, capable of performing exercise program, and without amino acid supplementation in the preceding six months were screened for eligibility. Exclusion criteria included renal or hepatic disease, carbohydrates intolerance or diabetes mellitus, corticosteroid use, restricted knee mobility (<5° extension or >120° flexion), persistent knee inflammation or pain (VAS ≥7/10), contraindications to isokinetic testing.

### Intervention

The experimental group received oral L-glutamine (0.75 g·kg ¹·day ¹), and control group isocaloric maltodextrin supplement **(Figure A)**. Supplements were provided in identical containers to blinding. Both groups followed an identical, standardized resistance training program **(Figure B, image 1)** in a sports medicine gym to ensure adherence, three times per week for six weeks, supervised by certified sports medicine specialists.

### Outcome

The main outcome was the peak torque of quadriceps and hamstring muscles, flexor/extensor ratio and limb symmetry index, measured at baseline and after six weeks using an isokinetic dynamometer (Biodex Medical Systems).

<u>Secondary outcomes</u> were lipid profile (triglycerides, HDL, LDL), liver enzymes (AST, ALT), and glucosa. Adverse events were monitored and recorded throughout the study period.

### Randomization

Participants were block randomized with 1:1 in blocks of six. The random sequence was computer-generated. An investigator not involved in participant recruitment or data collection was responsible for generating and safeguarding the allocation sequence in sealed opaque envelopes.

### Blinding

Both participants and researchers were blinded to group assignments. Supplements were prepared and distributed by personnel not involved in data collection or analysis.

### Sample Size Calculation

Sample size was estimated for anticipated 13% (SD 3%) difference in quadriceps strength gain between groups (19) with α=0.05 and 90% power, requiring 21 participants per group. Allowing for a 20% attrition, 25 participants were recruited per group.

### Statistical Analysis

Data were analyzed using Student’s t-test for normally distributed and the Mann-Whitney U test for non-normally distributed variables. Results were expressed as mean ± SD or median (IQR), respectively. A p-value of <0.05 was considered statistically significant. Analyses were conducted with JASP software.

### Ethical Considerations

The study was approved by the institutional ethics committee and complied with the Declaration of Helsinki. Informed consent was obtained from all participants. No conflicts of interest declared.

## Results

A total of 30 participants were enrolled, with 15 in each group. Data were available 30/30 (100%) of the participants at 6 weeks follow-up. Baseline characteristics are show in **Table 1**. No significant differences were observed between the L-glutamine (ExGln) and placebo (ExPla) groups in age, isokinetic parameters and anthropometric measures (p > 0.05). Biochemical variables were also comparable, except for C-LDL, which was higher in the ExPla group (p = 0.006). Primary outcomes were analyzed by Intention to treat; per protocol analysis included patients only who completed the study.

**Table 1.** Basal characteristics of sample under study.

| Variable | Group, Media (± DE) | | *P value* |
| --- | --- | --- | --- |
|  | ExGln<br>(n=15) | ExPla<br>(n=15) |  |
| Age, y | 34.0 ± 7.4 | 28.1 ± 7.5 | 0.081 |
| **Anthropometrics** |  |  |  |
| Mass, Kg | 81.5 ± 9.8 | 75.9 ± 15.4 | 0.525 |
| Height, cm | 173.6 ± 5.9 | 175.0 ± 8.2 | 0.697 |
| BMI, Kg/m<sup>2</sup> | 27.4 ± 2.9 | 26.9 ± 2.6 | 0.294 |
| Thigh circumference uninvolved limb, cm | 52.3 ± 3.0 | 53.3 ± 4.0 | 0.334 |
| Thigh circumference involved limb, cm | 49.4 ± 4.1 | 51.2 ± 4.1 | 0.790 |
| **Isokinetic** |  |  |  |
| Knee extensor strength, Nm |  |  |  |
| Uninvolved limb | 193.0 (174-211) | 193.0 (156-230) | 0.722 |
| Involved limb | 104.0 ± 34.9 | 108.0 ± 43.8 | 0.667 |
| Knee flexor strength, Nm |  |  |  |
| Uninvolved limb | 127.0 ± 13.8 | 122.0 ± 20.0 | 0.637 |
| Involved limb | 98.0 ± 17.4 | 98.0 ± 18.3 | 0.667 |
| Ratio flexor/extensor strength, % |  |  |  |
| Uninvolved limb | 65.6 ± 5.1 | 66.6 ± 11.3 | 0.377 |
| Involved limb | 104.0 ± 16 | 95.0 ± 58 | 0.334 |
| Knee extensor asymmetry index, % | 46.1 ± 16.3 | 38.5 ± 21.2 | 0.301 |
| Knee flexor asymmetry index, % | 19.0 ± 10.0 | 22.0 ± 15 | 0.151 |
| **Bioquímicas** |  |  |  |
| Glucose, mg/dl | 96.0 ± 7.5 | 93.0 ± 7.7 | 0.154 |
| Total Cholesterol, mg/dl | 179.0 ± 35.8 | 175.0 ± 32.9 | 0.580 |
| C-LDL, mg/dl | 101.0 ± 22 | 117.0 ± 24.2 | 0.006 |
| C-HDL, mg/dl | 42.0 ± 13.3 | 40.0 ± 3.5 | 0.525 |
| Triglycerides, mg/dl | 130.0 ± 37.7 | 115.0 ± 72.7 | 0.473 |
| AST, UI/L | 23.0 ± 5.2 | 21.0 ± 9.6 | 0.052 |
| ALT, UI/L | 29.0 ± 9.1 | 37.0 ± 8.0 | 0.101 |
| **Dietary** |  |  |  |
| Energy intake, kcal/d | 2321 ± 831 | 2480 ± 616 | 0.498 |
| Protein intake, gr/kg body weight | 1.2 ± 0.34 | 1.4 ± 0.27 | 0.377 |

Abbreviations: BMI, Body Mass Index; C-LDL, Cholesterol Low-Density Lipoprotein; C-HDL, Cholesterol High-Density Lipoprotein; AST, Aspartate Aminotransferase; ALT, Alanine Aminotransferase. p<0.05, U-Mann Whitney or $\chi^2$ test.

### Isokinetic Performance After 6 Weeks

**Table 2** presents the changes in isokinetic performance within and between groups after six weeks of intervention. In the ExGln group, knee-extension peak torque in the involved limb significantly (p = 0.001) as did the extensor-flexor ratio in that limb (p = 0.002). The ExPla group showed no meaningful changes in these measures. The Mann-Whitney U test showed a significant difference only for the extensor-flexor ratio of the involved limb (p = 0.045) but not for other isokinetic variables (p > 0.05). Regarding bilateral asymmetry, the ExGln group showed a clear reduction in extensor asymmetry after six weeks (p=000.1) and flexor asymmetry also decreased (0.018). In the ExPla group, only extensor asymmetry fell significantly (p=0.015). However, comparison between groups showed no significant differences in changes in extensor or flexor asymmetry (p=>0.05).

**Table 2.** Isokinetic differences within and between groups after 6 weeks.

| Variable | Grupo ExGln | | | Grupo ExPla | | | $\Delta$ medians<br>(IC 95%) | P<br>value** |
| --- | --- | --- | --- | --- | --- | --- | --- | --- |
|  | Basal | Final | *P<br>value | Basal | Final | *P<br>value |  |  |
| **PT Extensor (Nm)** |  |  |  |  |  |  |  |  |
| Uninvolved limb | 193 (171-198) | 199.6 (174-209) | 0.011 | 193 (153-212) | 196 (184-206) | 0.101 | 3.6 (-18-15) | 0.551 |
| Involved limb | 104 (84-125) | 163 (159-166) | 0.001 | 108 (88-157) | 161 (152-166) | 0.001 | 2.4 (-26-17) | 0.712 |
| **PT Flexor (Nm)** |  |  |  |  |  |  |  |  |
| Uninvolved limb | 127 (121-131) | 127 (125-137) | 0.807 | 122 (113-130) | 141 (132-149) | 0.001 | 9 (4.8-22.2) | 0.177 |
| Involved limb | 98 (91-107) | 126 (121-137) | 0.001 | 98 (77-110) | 120 (106-123) | 0.002 | 5 (-10-21.3) | 0.592 |
| **Ratio flex/Ext (%)** |  |  |  |  |  |  |  |  |
| Uninvolved limb | 64.5 (62-69) | 65 (59-72) | 0.798 | 61 (58.4-70) | 68 (62 - 70) | 0.495 | 2 (-0.51-9.5) | 0.714 |
| Involved limb | 104 (80 -109) | 83 (70 -102) | 0.002 | 80 (69.4-95.7) | 76 (65 - 80) | 0.378 | 5 (1.3-13) | 0.045 |
| **Bilateral asimmetry** |  |  |  |  |  |  |  |  |
| Extensores (%) | 41 (26.9-55.5) | 28.4 (13-29.6) | 0.001 | 38 (18-47) | 22 (13-29.6) | 0.015 | 6.4 (-3-37) | 0.198 |
| Flexores (%) | 20.8 (7.5-26.4) | 10.5 (0.80-18.1) | 0.018 | 19 (13-28) | 13 (9-15.2) | 0.361 | 3 (-8-29) | 0.082 |

PT= Peak Torque \*Wilcoxon Test, $p < 0.005$ ; \*\*U-Mann-Whitney, $p < 0.05$ .

### Biochemical Variables

Significant reductions were observed in the ExGln group for C-LDL (p = 0.006), C-HDL (p = 0.003), and triglycerides (p = 0.003) after six weeks of supplementation. In the ExPla group, only a significant reduction in triglycerides was observed (p = 0.009). Inter-group comparisons showed a significant difference for C-LDL (p = 0.031) but not for other biochemical parameters (p > 0.05).

**Table 3.** Biochemical variables between baseline and final evaluations.

| Variable | Grupo ExGln | | | Grupo ExPla | | | $\Delta$ medians<br>(IC 95%) | P<br>value** |
| --- | --- | --- | --- | --- | --- | --- | --- | --- |
|  | Basal | Final | *p*<br>value* | Basal | Final | *p*<br>value* |  |  |
| Glucose, mg/dl | 96.0 ± 7.5 | 98 (94-102) | 0.154 | 93.0 ± 7.7 | 90 (89-96) | 0.765 | 4.3 (-0.51-9.5) | 0.714 |
| Total Cholesterol, mg/dl | 179.0 ± 35.8 | 170 (141-191) | 0.580 | 175 (157-208) | 164 (150-191) | 0.009 | 4.9 (-16.3-26.4) | 0.294 |
| C-LDL, mg/dl | 101.0 ± 22 | 99 (87-107) | 0.006 | 117 (105-127) | 94 (91-117) | 0.005 | 8.3 (5.05-21.6) | 0.583 |
| C-HDL, mg/dl | 42.0 (34-55) | 53 (47-54) | 0.003 | 40.0 ± 3.5 | 42 (39-54) | 0.178 | 5.9 (3.2-12.6) | 0.820 |
| Triglycerides, mg/dl | 130 (77-178) | 88 (70-108) | 0.003 | 115.0 ± 72.7 | 122 (92-149) | 0.642 | 22.3 (24-93.1) | 0.604 |
| AST, UI/L | 23.0 ± 5.2 | 24 (21-29) | 0.052 | 21.0 ± 19.6 | 21 (19-23) | 0.905 | 2.6 (-1.09-6.42) | 0.332 |
| ALT, UI/L | 29.0 ± 9.1 | 35 (32-49) | 0.101 | 37.0 ± 18.0 | 33 (29-37) | 0.710 | 9.2 (-72-21) | 0.057 |

\*Wilcoxon Test, $p < 0.005$ ; \*\*U-Mann-Whitney, $p < 0.05$ .

### Summary of Findings

Overall, the results suggest that glutamine supplementation combined with a standardized strength training program may enhance isokinetic performance, reduce bilateral asymmetry, and improve certain biochemical parameters in patients undergoing ACL reconstruction. However, the inter-group differences for most outcomes did not reach statistical significance, indicating that further research may be needed to confirm these findings.

## Discussion

### Principal Findings

This study evaluated the effects of oral glutamine supplementation combined with a standardized strength training program on isokinetic performance, bilateral asymmetry, and biochemical variables in male patients undergoing anterior cruciate ligament reconstruction (ACLR). The results suggest that glutamine supplementation may enhance knee extensor strength of the involved limb, reduce bilateral asymmetry. However, despite these within-group improvements, inter-group comparisons revealed limited significant differences, indicating that the observed benefits might partially be due to the synergistic effects of exercise and supplementation rather than supplementation alone. (12, 17, 19).

### Comparison with Previous Studies

The observed reduction in knee extensor asymmetry and improvements in peak torque in the ExGln group align with previous studies demonstrating the anabolic and anti-catabolic effects of glutamine in muscle recovery post-ACLR (18, 27). Glutamine’s role in supporting protein synthesis, immune modulation, and antioxidative defense may explain the enhanced isokinetic performance observed (28). The reductions in C-LDL and triglycerides levels further corroborate findings suggesting that exercise training can modulate lipid metabolism, likely due to energy expenditure and effects on liver enzymes (29). Additionally, glutamine may contribute by enhancing glutathione synthesis and reduction of oxidative stress (30).

In contrast, the limited inter-group differences imply that the benefits of glutamine may be more pronounced when combined with a structured rehabilitation program, emphasizing the importance of exercise-induced muscle hypertrophy and neuromuscular adaptation (**Table 2**) and strength training alone can significantly reduce bilateral asymmetry, suggesting that the effects observed may not be attributable solely to glutamine supplementation (12, 22).

### Clinical Implications

The findings underscore the potential role of nutritional supplementation as an adjunct to standard ACLR rehabilitation. The improvements in C-LDL and triglycerides indicate cardiometabolic benefits of exercise, which may be relevant for patients with metabolic syndrome or dyslipidemia (30). The reduction in bilateral asymmetry in the ExGln group may also support better functional recovery and lower reinjury risk, on line with clinical guidelines emphasizing restoration of muscular symmetry (12, 31).

However, the absence of significant inter-group differences for most variables raises questions about the cost-effectiveness and clinical significance of glutamine supplementation (18). It may be more beneficial in catabolic states or in patients with significant deficits in protein intake rather than as a standard adjunct for all ACLR patients.

### Limitations

This study has several limitations that warrant consideration. First, the sample size was relatively small, potentially limiting the statistical power to detect differences between groups. Second, the study was conducted exclusively in male patients, limiting the generalizability of findings to female populations, who may exhibit different responses due to variations in hormonal status and muscle metabolism (32). Third, we did not assess Arthrogenic Muscle Inhibition, which often follows ACL reconstruction and is considered a barrier to restoring muscle strength.

Additionally, the use of isokinetic testing to assess muscle strength, while objective, may not fully reflect functional outcomes such as return-to-sport or quality of life, which are critical in evaluating the success of ACLR rehabilitation (29). Future studies with larger, more diverse populations and extended follow-up durations are needed to validate these findings.

### Future Directions

Further research should explore the potential of glutamine supplementation in catabolic states or in patients with malnutrition or sarcopenia undergoing ACLR. Investigating the effects of combined amino acid formulations that include branched-chain amino acids (BCAAs) alongside glutamine could provide insights into optimizing nutritional strategies for muscle recovery (33). Moreover, randomized trials assessing functional outcomes such as agility, balance, and return-to-sport timelines would offer a more comprehensive understanding of the clinical benefits of glutamine supplementation.

## Conclusion

The findings of this study suggest that oral glutamine supplementation, when combined with a standardized strength training program, can enhance isokinetic performance and reduce bilateral asymmetry in male patients following anterior cruciate ligament reconstruction (ACLR). The observed improvements in knee extensor strength, C-LDL, and triglycerides in the supplementation group highlight the potential of glutamine as an effective adjunct to rehabilitation protocols. However, the limited inter-group differences imply that the benefits may predominantly stem from the structured exercise regimen rather than glutamine supplementation alone.

The results underscore the importance of comprehensive rehabilitation programs that integrate nutritional strategies with targeted strength training to optimize muscle recovery and functional outcomes post-ACLR. The significant reduction in bilateral asymmetry suggests that glutamine supplementation might aid in restoring neuromuscular balance, potentially lowering the risk of reinjury and enhancing return-to-sport timelines. Moreover, the improvements in lipid profiles may offer additional cardiometabolic benefits, which could be especially relevant for ACLR patients with pre-existing metabolic risk factors.

Despite these promising findings, the small sample size and male-only cohort the generalizability of the results. Larger and more diverse populations and extended follow-up durations are needed to confirm these findings and to clarify the clinical utility of glutamine supplementation in ACLR rehabilitation. Future studies should also examine combined amino acid strategies and their impact on functional outcomes such as balance, agility, and quality of life.

In conclusion, while glutamine supplementation shows potential as an adjunct to traditional rehabilitation and exercise protocols, its routine use post-ACLR should be approached with caution until further evidence is available. Comprehensive, long-term clinical trials are warranted to establish definitive evidence-based recommendations for the integration of nutritional supplementation into ACLR exercise protocols.

## Data Availability

All data produced in the present work are contained in the manuscript

---

**Figure A.**
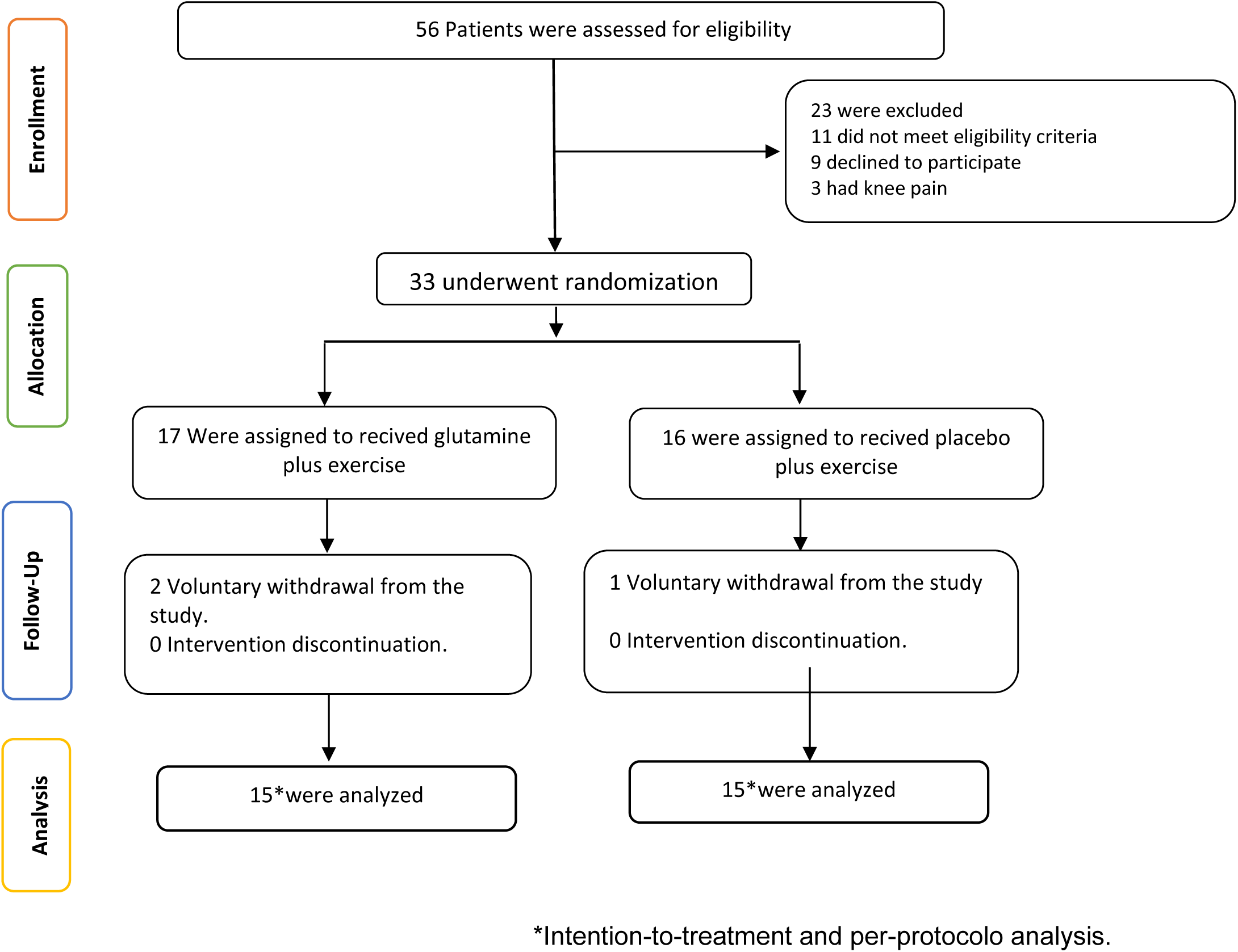
**CONSORT Flow of Clinical Control Trial.**

**Figure B.**
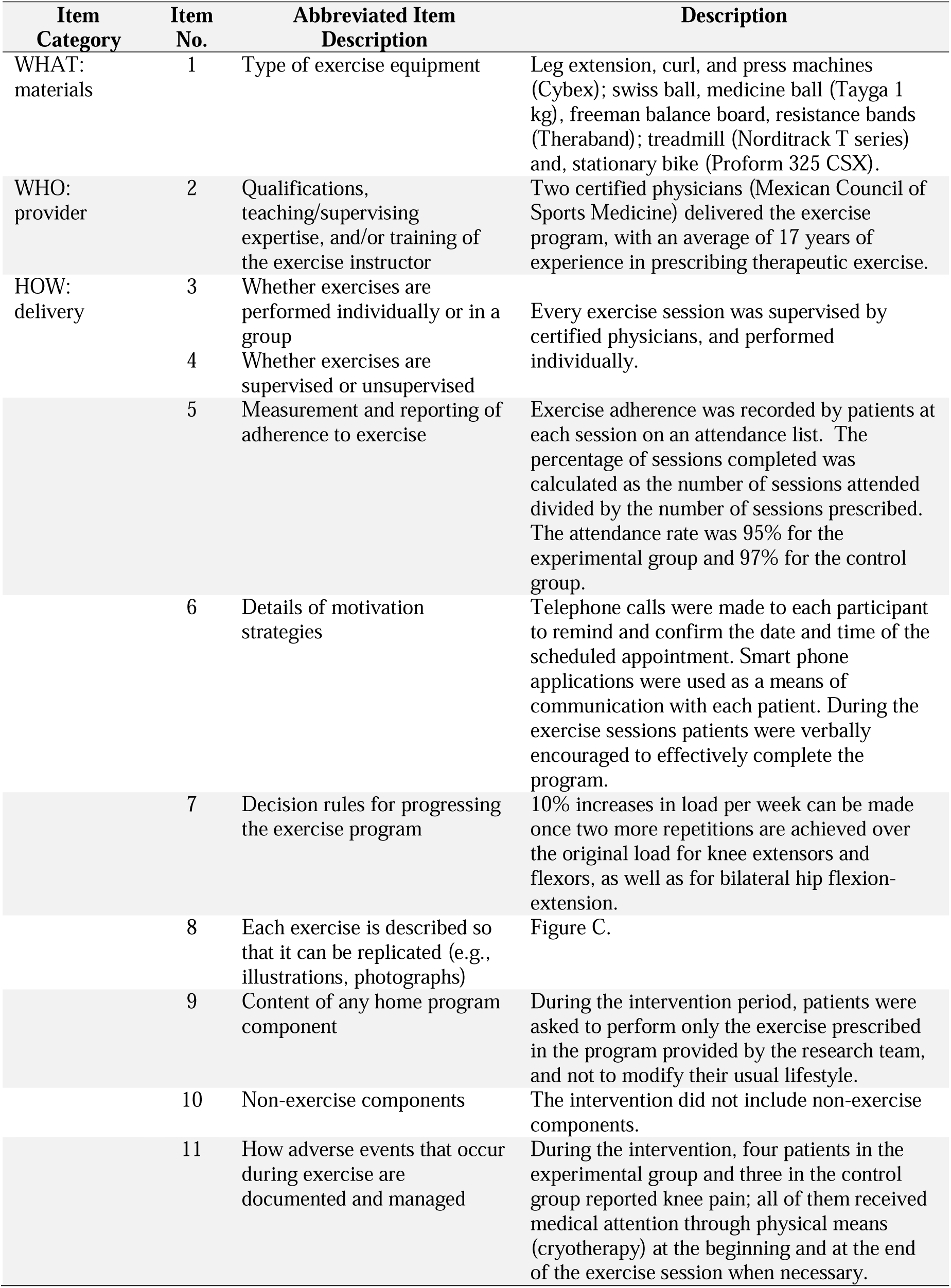
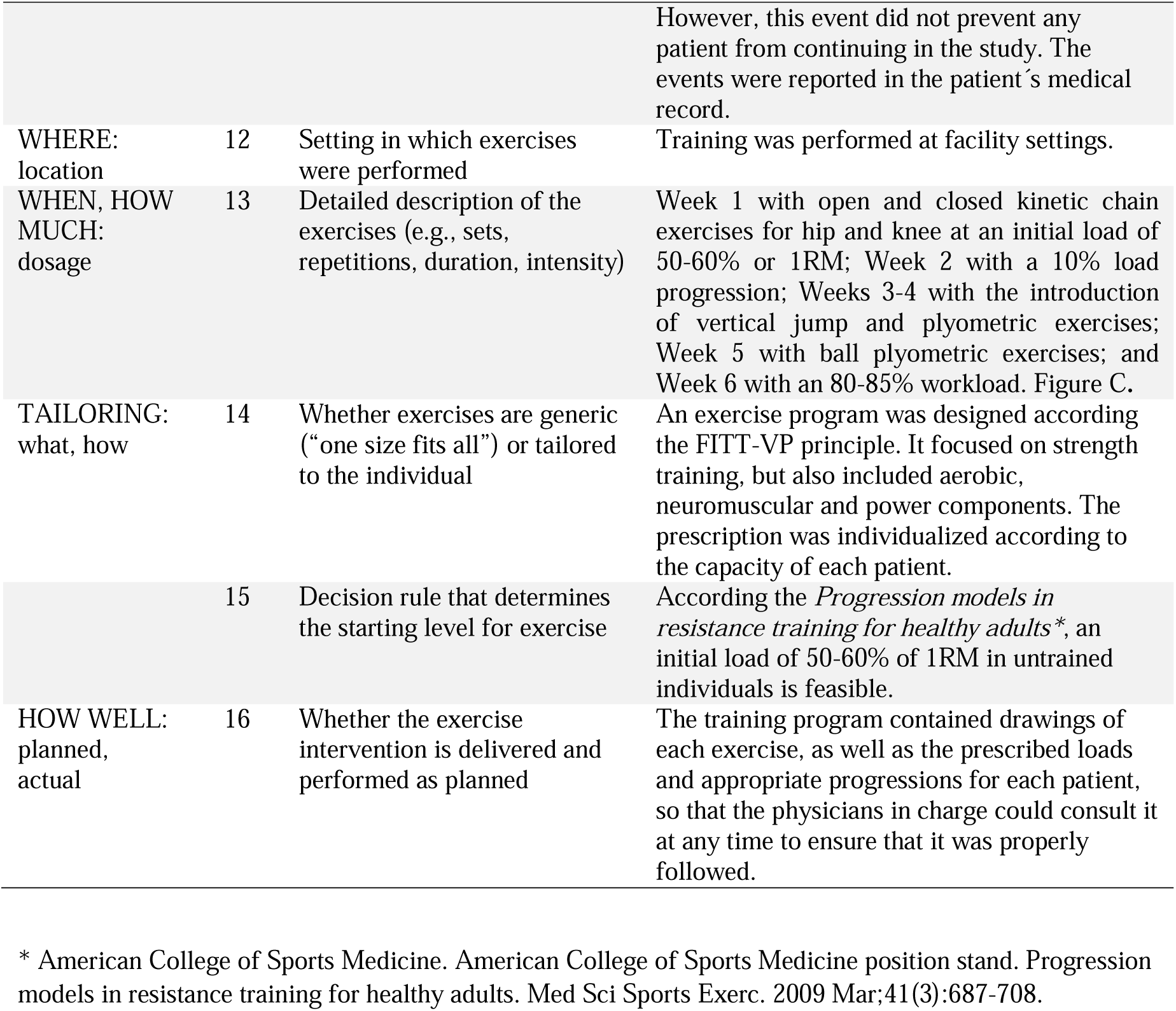
**Exercise Reporting Template.**

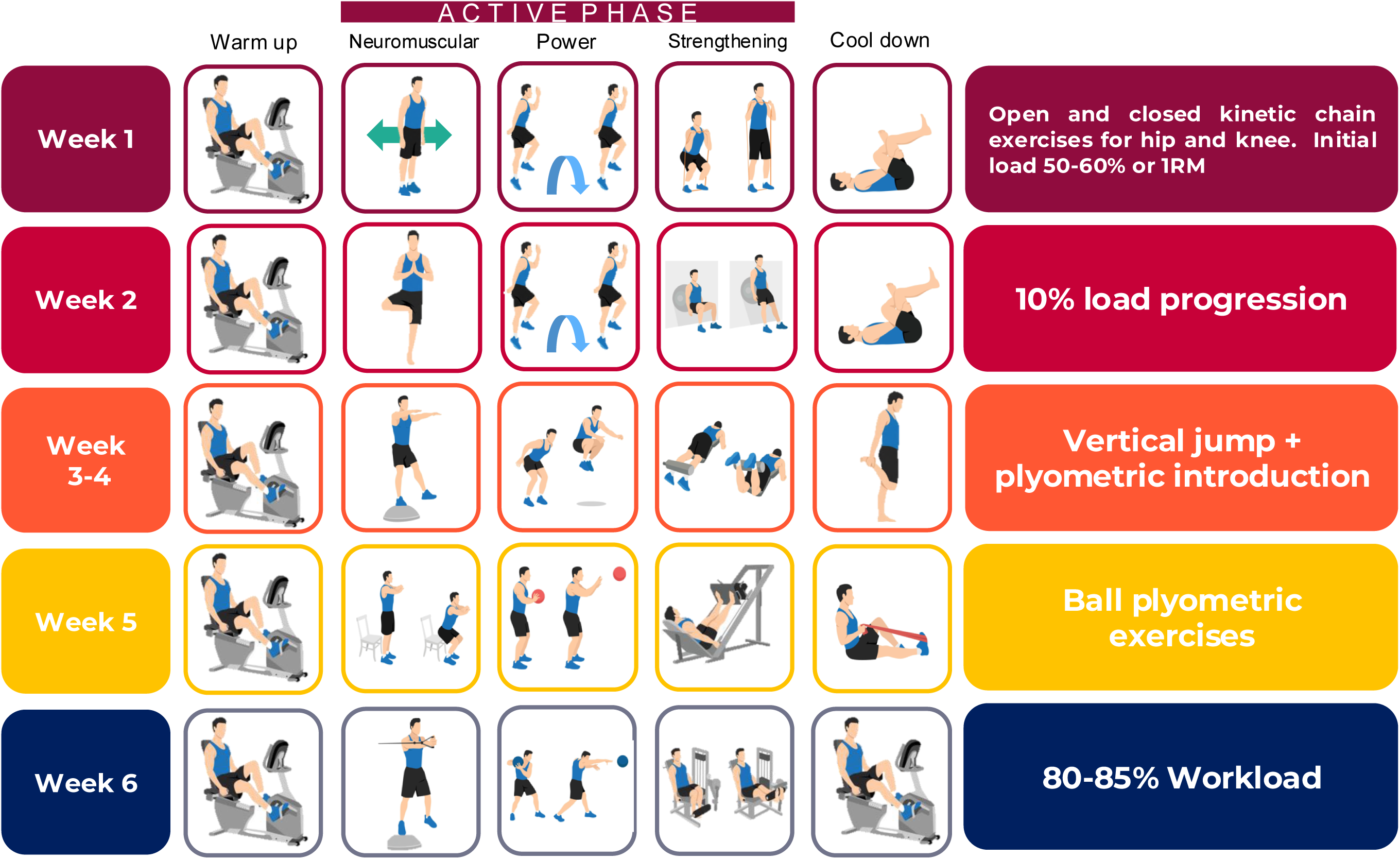

## References

1. Siegel L, Vandenakker-Albanese C, Siegel D. Anterior cruciate ligament injuries: anatomy, physiology, biomechanics, and management. Clin J Sport Med. 2012;22(4):349–55.

2. Cimino F, Volk BS, Setter D. Anterior cruciate ligament injury: diagnosis, management, and prevention. Am Fam Physician. 2010;82(8):917–22.

3. Moses B, Orchard J, Orchard J. Systematic review: Annual incidence of ACL injury and surgery in various populations. Res Sports Med. 2012;20(3-4):157–79.

4. Webster KE, Hewett TE. Anterior Cruciate Ligament Injury and Knee Osteoarthritis: An Umbrella Systematic Review and Meta-analysis. Clin J Sport Med. 2022;32(2):145–52.

5. Nawasreh Z, Logerstedt D, Cummer K, Axe MJ, Risberg MA, Snyder-Mackler L. Do Patients Failing Return-to-Activity Criteria at 6 Months After Anterior Cruciate Ligament Reconstruction Continue Demonstrating Deficits at 2 Years? Am J Sports Med. 2017;45(5):1037–48.

6. Hsiao SF, Chou PH, Hsu HC, Lue YJ. Changes of muscle mechanics associated with anterior cruciate ligament deficiency and reconstruction. J Strength Cond Res. 2014;28(2):390–400.

7. Wang LJ, Zeng N, Yan ZP, Li JT, Ni GX. Post-traumatic osteoarthritis following ACL injury. Arthritis Res Ther. 2020;22(1):57.

8. Kuenze CM, Hertel J, Weltman A, Diduch D, Saliba SA, Hart JM. Persistent neuromuscular and corticomotor quadriceps asymmetry after anterior cruciate ligament reconstruction. J Athl Train. 2015;50(3):303–12.

9. Delfino GB, Peviani SM, Durigan JL, Russo TL, Baptista IL, Ferretti M, et al. Quadriceps muscle atrophy after anterior cruciate ligament transection involves increased mRNA levels of atrogin-1, muscle ring finger 1, and myostatin. Am J Phys Med Rehabil. 2013;92(5):411–9.

10. Dos Anjos T, Gabriel F, Vieira TD, Hopper GP, Sonnery-Cottet B. Neuromotor Treatment of Arthrogenic Muscle Inhibition After Knee Injury or Surgery. Sports Health. 2024;16(3):383–9.

11. Kruse LM, Gray B, Wright RW. Rehabilitation after anterior cruciate ligament reconstruction: a systematic review. J Bone Joint Surg Am. 2012;94(19):1737–48.

12. Glattke KE, Tummala SV, Chhabra A. Anterior Cruciate Ligament Reconstruction Recovery and Rehabilitation: A Systematic Review. J Bone Joint Surg Am. 2022;104(8):739–54.

13. Lapaeva AG, Tabakov RS, Tabakov SE, Miroshnikov AB, Smolensky AV. [Effect of dietary supplements and whey protein on muscle mass and strength of the operated limb after anterior cruciate ligament reconstruction: a systematic review]. Vopr Pitan. 2023;92(2):87–96.

14. Pasiakos SM, McLellan TM, Lieberman HR. The effects of protein supplements on muscle mass, strength, and aerobic and anaerobic power in healthy adults: a systematic review. Sports Med. 2015;45(1):111–31.

15. Eichner ER. Glutamine supplementation: overstaying its welcome. Curr Sports Med Rep. 2013;12(4):211–2.

16. Agostini F, Biolo G. Effect of physical activity on glutamine metabolism. Curr Opin Clin Nutr Metab Care. 2010;13(1):58–64.

17. Ramezani Ahmadi A, Rayyani E, Bahreini M, Mansoori A. The effect of glutamine supplementation on athletic performance, body composition, and immune function: A systematic review and a meta-analysis of clinical trials. Clin Nutr. 2019;38(3):1076–91.

18. Cruzat V, Macedo Rogero M, Noel Keane K, Curi R, Newsholme P. Glutamine: Metabolism and Immune Function, Supplementation and Clinical Translation. Nutrients. 2018;10(11).

19. Piattoly T, Parish T, Welsch M. L-glutamine supplementation: Effects on endurance, power and recovery. Current Topics in Nutraceutical Research. 2013;11:55–62.

20. Mok E, Constantin B, Favreau F, Neveux N, Magaud C, Delwail A, Hankard R. l-Glutamine administration reduces oxidized glutathione and MAP kinase signaling in dystrophic muscle of mdx mice. Pediatr Res. 2008;63(3):268–73.

21. dos Santos RV, Caperuto EC, de Mello MT, Batista ML, Jr., Rosa LF. Effect of exercise on glutamine synthesis and transport in skeletal muscle from rats. Clin Exp Pharmacol Physiol. 2009;36(8):770–5.

22. McGlory C, Devries MC, Phillips SM. Skeletal muscle and resistance exercise training; the role of protein synthesis in recovery and remodeling. J Appl Physiol (1985). 2017;122(3):541–8.

23. Hernández Valencia SE, Méndez Sánchez L, Clark P, Moreno Altamirano L, Mejía Aranguré JM. [GLUTAMINE AS AN AID IN THE RECOVERY OF MUSCLE STRENGTH: SYSTEMATIC REVIEW OF LITERATURE]. Nutr Hosp. 2015;32(4):1443–53.

24. Legault Z, Bagnall N, Kimmerly DS. The Influence of Oral L-Glutamine Supplementation on Muscle Strength Recovery and Soreness Following Unilateral Knee Extension Eccentric Exercise. Int J Sport Nutr Exerc Metab. 2015;25(5):417–26.

25. Holm L, Esmarck B, Mizuno M, Hansen H, Suetta C, Hölmich P, et al. The effect of protein and carbohydrate supplementation on strength training outcome of rehabilitation in ACL patients. J Orthop Res. 2006;24(11):2114–23.

26. Schulz KF, Altman DG, Moher D. CONSORT 2010 Statement: updated guidelines for reporting parallel group randomised trials. BMC Med. 2010;8:18.

27. Rodrigues Junior CF, Murata GM, Gerlinger-Romero F, Nachbar RT, Marzuca-Nassr GN, Gorjão R, et al. Changes in Skeletal Muscle Protein Metabolism Signaling Induced by Glutamine Supplementation and Exercise. Nutrients. 2023;15(22).

28. Córdova-Martínez A, Caballero-García A, Bello HJ, Pérez-Valdecantos D, Roche E. Effect of Glutamine Supplementation on Muscular Damage Biomarkers in Professional Basketball Players. Nutrients. 2021;13(6).

29. Smart NA, Downes D, van der Touw T, Hada S, Dieberg G, Pearson MJ, et al. The Effect of Exercise Training on Blood Lipids: A Systematic Review and Meta-analysis. Sports Med. 2025;55(1):67–78.

30. Leite JSM, Vilas-Boas EA, Takahashi HK, Munhoz AC, Araújo LCC, Carvalho CR, et al. Liver lipid metabolism, oxidative stress, and inflammation in glutamine-supplemented ob/ob mice. J Nutr Biochem. 2025;138:109842.

31. Kotsifaki R, Korakakis V, King E, Barbosa O, Maree D, Pantouveris M, et al. Aspetar clinical practice guideline on rehabilitation after anterior cruciate ligament reconstruction. Br J Sports Med. 2023;57(9):500–14.

32. Zhao D, Liang GH, Pan JK, Zeng LF, Luo MH, Huang HT, et al. Risk factors for postoperative surgical site infections after anterior cruciate ligament reconstruction: a systematic review and meta-analysis. Br J Sports Med. 2023;57(2):118–28.

33. Greif DN, Emerson CP, Allegra P, Arizpe A, Mansour KL, Cade WH, 2nd, Baraga MG. Supplement Use in Patients Undergoing Anterior Cruciate Ligament Reconstruction: A Systematic Review. Arthroscopy. 2020;36(9):2537–49.

